# Using a co-design approach to develop aquatic reactive balance training for fall prevention

**DOI:** 10.64898/2026.03.07.26347842

**Authors:** Anna Ogonowska-Slodownik, Júlia O. Faria, Shanuga Thavarajah, Karina Pacholczyk, Birgit Blain, Myra Wiener, Jane Walker, Avril Mansfield, Soo Chan Carusone

## Abstract

**Background:** Falls are a major health concern among older adults, leading to injury, reduced independence, and increased healthcare cost. Reactive balance training can reduce fall risk, but barriers such as fear, joint discomfort, and harness burden limit its use - barriers that aquatic training may help overcome.

**Objective:** To design an aquatic reactive balance training (AquaReBal) program for older adults, integrating end-user perspectives to enhance safety, accessibility, and engagement.

**Methods:** Using a participatory design approach, we engaged older adult partners, physiotherapists, and researchers in iterative phases including literature review, stakeholder consultations, practical pool sessions, and feedback meetings. Data were collected through online meetings, surveys, and real-time observations, following the Guidance for Reporting Involvement of Patients and the Public 2 framework.

**Results:** Three older adult partners and a multidisciplinary team co-designed the AquaReBal protocol through two participatory design sessions, one practical pool session, and two internal team sessions. Key recommendations from partners included using a vest instead of a hip belt for perturbations, addressing pool depth visibility, and creating an introductory package with practical information for participants. Partners emphasized safety, instructor support, and social engagement as critical for adherence and satisfaction.

**Conclusion:** The co-design process enabled the development of an AquaReBal protocol tailored to older adults’ needs and preferences, demonstrating potential for broader implementation.

## Introduction

Falls are a major health concern among older adults, leading to injury, reduced independence, and increased healthcare cost [1]. One of the current fall prevention methods for adults is reactive balance training (RBT), where individuals experience an intentional loss of balance and must complete steps to regain it to avoid the fall [2]. While land-based RBT has been shown to reduce fall risk, its accessibility is limited for some older adults due to fear of falling or injury, joint discomfort, and the physical burden of harness use [3]. Given the effectiveness of RBT, addressing its barriers is crucial to maximizing its reach and impact.

One method to overcome these barriers with land-based RBT is to conduct the training in an aquatic environment. Water offers unique advantages, such as buoyancy and hydrostatic pressure, which can be used in RBT [4]. Water properties may enhance comfort and confidence. While these theoretical benefits exist, only one study to date has tested this concept in the water [5], but it did not assess reactive balance control or falls.

Co-design has been defined as collaboration between researchers, end-users, and stakeholders to iteratively create tailored interventions [6]. Involving older adults in research allows them to develop new skills and fosters an inclusive research process [7]. However, it remains underutilized in older adult population [8, 9]. Co-designing interventions with older adults could help enhance their participation in programs specifically designed for them [10].

Involving stakeholders in the design process helps shape the program to to shape the program to better meet the needs of end-users. In this study, we define Patient and Public Involvement (PPI) as a partnership between researchers and older adults, where both groups work together to make decisions, learn from each other, and co-create the program.

Incorporating PPI in our study is grounded in participatory health research theories, which support involving end-users in developing solutions. By including stakeholders’ perspectives from the start, we aimed to build a program that reflects their real-world experiences while improving accessibility, especially for those facing barriers with traditional land-based balance training. This study adds to the growing research on participatory and user-centered design by highlighting the active role older adults can play, not only in creating interventions but also in disseminating results.

The aim of this paper is to describe the co-design of an aquatic reactive balance training (AquaReBal) program for older adults and the impact of engaging older adults with relevant lived experience.

## Materials and methods

### Study Design

Our co-design study employed different methods to generate insights and design the final AquaReBal program. Collective decision-making and knowledge exchange among the involved stakeholders played a key role throughout this process. We used the Guidance for Reporting Involvement of Patients and the Public 2 (GRIPP2) to report the patient and public involvement activities in this study[11]. This study also adopted the principles of the Participatory Action Research (PAR) approach. PAR emphasizes the active involvement of stakeholders through ongoing collaboration with researchers, following an iterative cycle of fieldwork or practice, reflection, planning, research, and action [12, 13], aligning closely with co-design methodologies [14]. The goal was to enable the intended end users of the intervention to make decisions about its implementation based on their needs and expectations, thereby achieving outcomes that reflect their perspectives and their lived experiences [15].

The suggestions gathered in each session guided the development of AquaReBal program by identifying facilitators and barriers to practice adherence, and helped us understand the level of difficulty perceived by the older adults for the intended exercises [16]. This is particularly relevant given that RBT is a challenging type of balance training in which participants are repeatedly exposed to balance perturbations and perform balance reactions to avoid falls [2]. The study was approved by the University Health Network Research Ethics Board (study ID: 24-6146).

### Participants

The research team was created in February 2025 included five physiotherapists (three specialized in aquatic therapy, two in land-based RBT), a researcher with extensive RBT experience, and two master’s students in translational research. Three older adult partners were involved - three women over the age of 65 years. Two of them had previously participated in a randomized controlled trial of land-based reactive balance training, although their group allocation was not known. One of the partners had experience in exercising in an aquatic environment, as she regularly attended weekly aquatic exercise sessions. One partner had no experience in aquatic or RBT, but was participating in a fall prevention exercise group, aligning with the protocol’s goals of reducing fall incidence. All of the partners experienced a fall in the last year.

### Co-design sessions

The process followed a series of steps: initial engagement, relationship building, identification of concerns, participatory action, implementation, and subsequent reflection and evaluation [17]. Although the steps are described as separate, they often overlap and evolve dynamically throughout the process [18]. The process was structured into multiple iterative phases involving both the research team and partners (Fig. 1). All meetings aside from the practical sessions in the pool were conducted online through MS Teams.

**Fig 1.**
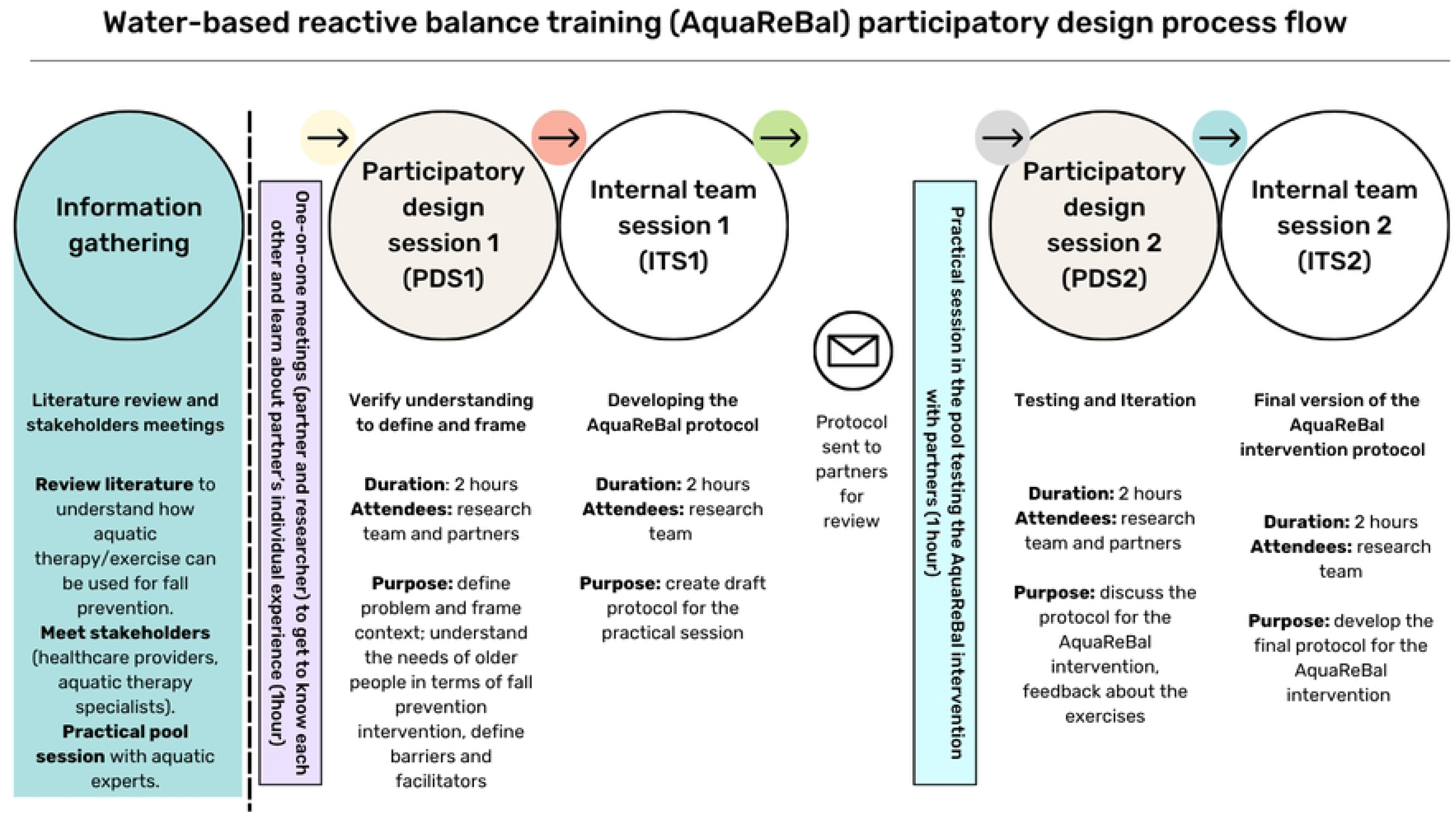
Co-design process

Initially, an information gathering phase was conducted, including a literature review to explore the use of aquatic exercise in fall prevention and stakeholder meetings with healthcare professionals and aquatic specialists to contextualize the intervention. Next, we recruited the older adult partners and individual one-on-one meetings were held between each partner and the principal researcher to understand their personal experiences with falls, aquatic exercise, and balance training.

Following this, the first participatory design session (PDS1) was held with the research team and older adult partners. This group session aimed to frame the problem collaboratively, explore the needs and preferences of older adults regarding RBT in water, and identify potential barriers and facilitators to the intervention. Insights from this session were synthesized and discussed during the first internal team session (ITS1) meeting involving researchers, from which a protocol for the practical session was created. One week prior to the practical session in the pool, the protocol was shared with the partners for review. Practical sessions were conducted individually with each partner in the pool to test the proposed exercises. They enabled the research team to evaluate the feasibility, comfort, safety, and challenge level of the activities and ways of delivering perturbations. Based on this experience, a second participatory design session (PDS2) was conducted involving both researchers and partners. During this session, feedback from the practical session was discussed, and suggestions for modifying the exercises were collected and incorporated through discussion. A second internal team session (ITS2) was conducted, during which the research team finalized the AquaReBal protocol, integrating suggestions from all prior phases.

All online sessions were audio-recorded and documented with notes to support analysis and ensure transparency and rigor in the co-design process. This multi-step, collaborative methodology aligns with recommendations for involving end users in the design of health interventions to enhance their relevance, usability, and potential for implementation [19].

## Results

In this section, we present the multi-step co-design process used to develop the AquaReBal program. After each step, feedback was gathered and reviewed in detail. The insights and recommendations informed specific modifications. As a result, the project evolved dynamically, reflecting lessons learned throughout its development. The results are organized according to the sequence of activities conducted.

### Information gathering

First, a comprehensive literature review was performed to synthesize current evidence on aquatic therapy, RBT, and their potential integration. There were only two other studies that used perturbations in the water as a part of the program. An online meeting was held to discuss key principles of aquatic-based exercise and the application of RBT to water environment with international aquatic therapy experts from US, Canada, Australia and Europe (n=7). This was followed by a practical session in the pool with the aquatic therapy experts (n=4) to directly observe and experience the properties of water that could influence RBT. Consultations were conducted with experts in land-based RBT, who provided insights into the key mechanisms, safety considerations, and progressions involved in delivering interventions. Importantly, the AquaReBal program is conceptually based on the Toronto Perturbation-Based Balance Training framework, which emphasizes task-specific, progressively challenging perturbations to improve reactive balance control and reduce fall risk. Collectively, these steps formed the foundation to conduct the co-design of AquaReBal, ensuring that the intervention was evidence-informed and tailored to aquatic environment.

### One-on-one meetings

To get to know the older adult partners (n=3) and build trust, one-on-one meetings were conducted online by the principal researcher with each partner. The conversation included the purpose of the project, steps involved, and their role; their experiences with physical activity, including aquatic exercise; and their perceptions of balance, falls, and reactive balance training on land. See Figure 2 for key themes and quotes from these initial one-on-one meetings with partners (Fig. 2).

**Fig 2.**
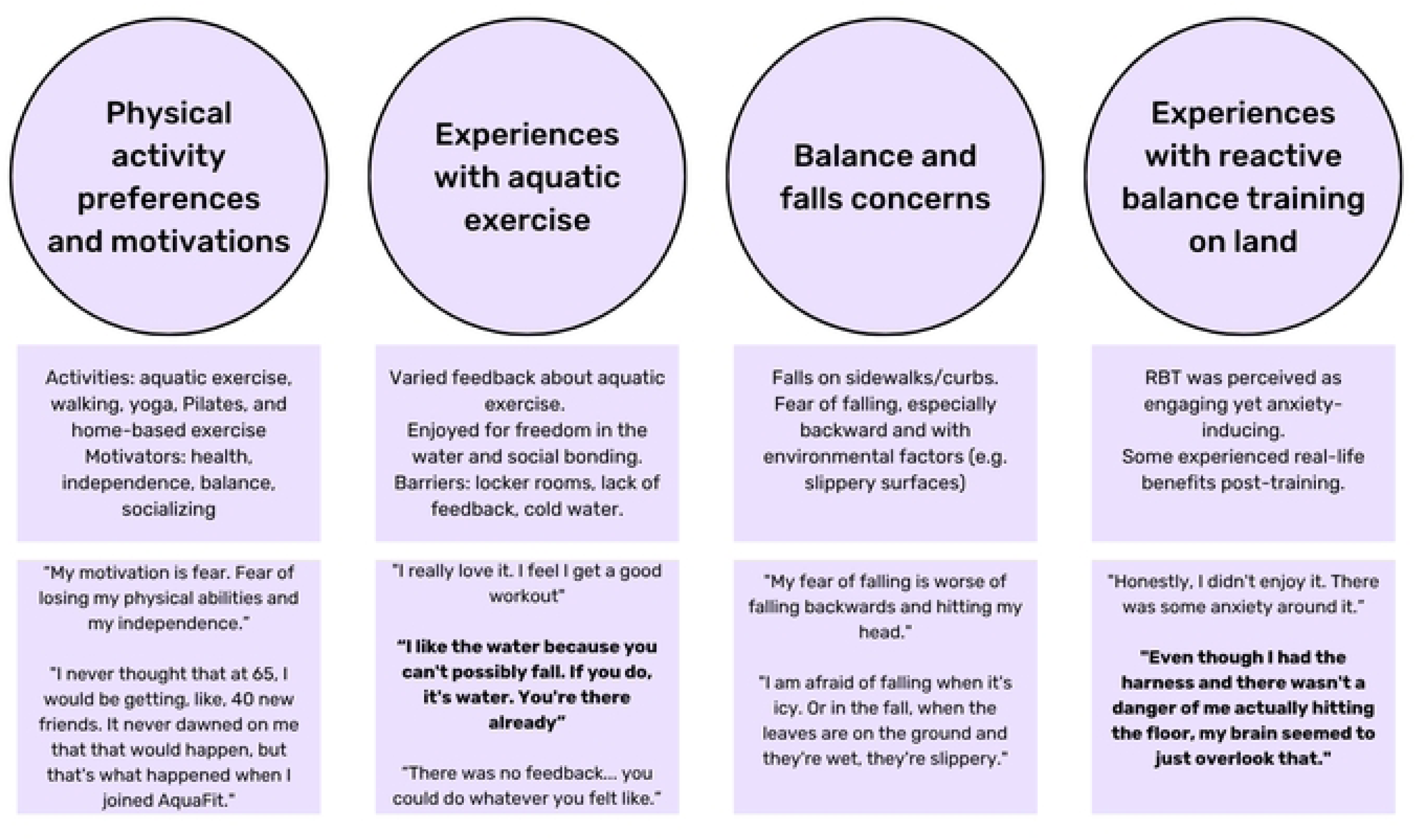
Lived experience context from the one-on-one meetings. Quotes bolded are to highlight the feelings regarding water environments and the experiences with land-based RBT.

Partners engaged in a range of physical activities and motivations included maintaining health, preventing physical decline, improving balance, and fostering social connections. Experiences with aquatic exercise were mixed. While some partners described it as enjoyable and socially engaging others reported barriers such as discomfort in locker rooms, lack of feedback, and cold or slippery environments. Concerns about balance and falling were prevalent, often tied to past experiences with falls. Fear of falling, particularly backward, was a recurring theme. Experiences with land-based RBT were diverse. One partner found the unpredictability stimulating, while the other experienced anxiety despite safety equipment. Notably, one partner reported improved balance confidence and responses to slips after land-based RBT. These insights directly shaped the development of AquaReBal, emphasizing safety, progressive challenge, and personalized support within an aquatic setting.

### Participatory Design Session 1

The first participatory design session (PDS1) was conducted online, followed by a post-session anonymous survey to gather feedback on the process of meeting. The meeting was attended by the principal researcher, two master’s students, three partners, one land-based RBT expert, and one aquatic therapy expert (n=8). The meeting generated insights about the intervention as well as ideas to create a more inclusive and welcoming program, including the development of an ‘Introductory package’ for participants. Ideas brought up throughout the meeting were captured using a Miro board, for example when discussing motivators, facilitators and barriers to aquatic program participation.

Motivations for taking part in aquatic interventions in general were mainly in the areas of health and functional well-being, social engagement, experience of being in the water environment and the influence of the instructors. Partners emphasized the important role of the instructor. According to them, a strong connection with the instructor enhances motivation, enjoyment, and continued participation.

> *“If you enjoy the people that you’re with, like the instructor and your co-participants, that it could lift your mood as well.” (Partner)*

> *“I think the instructor is very important because they’re the ones that are motivating you to work harder, and if you don’t have that connection you won’t wanna go because it’s no longer fun.” (Partner)*

Barriers discussed focused on five main categories that can limit participation in aquatic exercise (Fig 3): health and physical safety concerns, comfort and confidence in the water, program design and suitability, practical and logistical barriers, and financial barriers. One partner mentioned how past negative experiences, even from early life, can have a lasting impact on willingness to engage in water-based programs.

**Fig 3.**
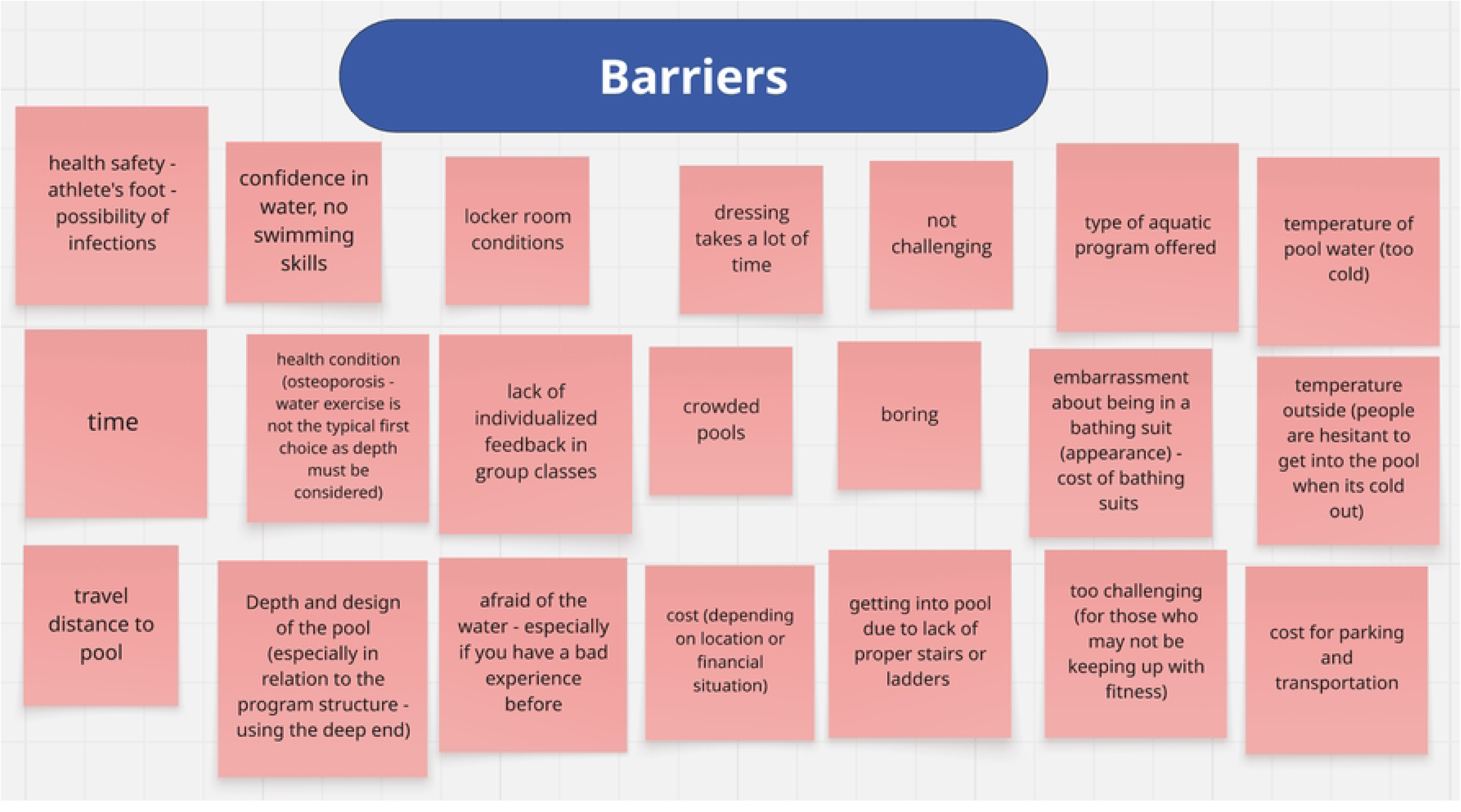
Miro board with the barriers to aquatic exercise discussed in the first participatory design session.

The PDS1 meeting was evaluated in an anonymous way with a post-meeting survey (n=7) to gather feedback. One person did not complete the survey. This was used as another way to gather feedback from meeting attendees to gauge how they felt about the meeting and areas of improvement for future meetings. The respondents were overall very satisfied with the meeting based on the scoring (4.8/5). They found videos of water exercises especially helpful and engaging. The use of the Miro board format was brought up a few times. By some, it was seen as a valuable tool that helped ensure everyone was on the same page, while others found it a bit distracting since it did not work as smoothly as hoped. Respondents enjoyed meeting everyone involved in the project, and they found the input from other stakeholders meaningful, especially when it was grounded in their clinical experience and connected to participants’ real-world needs. Everyone reported that their ideas were heard during the session and brainstorming was seen as a valuable part of the meeting. One person recommended including time for final reflections at the end of future meetings to allow everyone to reflect on how they felt and share anything they learned during the session.

### Internal Team Session 1

The first internal team session included research team members (n=5) and was conducted following PDS1 to review and synthesize feedback from older adult partners regarding the initial program concepts, session structure, and motivational strategies. The research team discussed insights gathered through the Miro board, including barriers to aquatic exercise, preferences for instructor support, and ideas for progress tracking. Decisions were made to refine the ‘Introductory package’, clarify session instructions, and adjust proposed exercises to better accommodate comfort, safety, and engagement. This session ensured that partner feedback was systematically incorporated into the program design before the practical pool session. Protocol for the pool session was created and sent to the partners.

### Practical pool sessions

Individual practical sessions were carried out in a pool environment to test and refine potential exercises and equipment for the AquaReBal program with three partners involved in our project. The practical sessions were conducted by two physiotherapists (one in the water leading the session and one on land taking notes and registering what was done). The sessions took place at the Toronto Rehabilitation Institute. The pool is situated on the second floor, with easy access via elevators and stairs. Individual changing rooms are available, ensuring privacy, a factor previously mentioned as a barrier during the first participatory design session by one partner.

During the practical pool session, partners trialed a range of aquatic reactive balance exercises designed to target task-specific perturbations and allow for progressive challenge. The activities included walking tasks (forward, backward, tandem walking, and walking in place), performed at different depths and, in some cases, with eyes closed or modified arm positions to increase task complexity. Lean-and-release was performed in forward, backward, and lateral directions, both in shallow and deeper areas of the pool. Additional perturbations were delivered using equipment, including multidirectional pulls with a waist belt during standing, walking, and rapid stepping, as well as manual pushes and pulls.

Each exercise was initially performed at a comfortable level and was then adapted to increase difficulty, for example, closing the eyes, taking longer steps, narrowing the base of support, or applying additional perturbations. The training progression was tailored to each individual’s abilities and level of confidence in the water. After each exercise, perceived intensity was recorded using the Balance Intensity Scale (BIS) [20].

### Participatory Design Session 2

The meeting was attended by the principal researcher, two students, three partners, one land-based RBT expert, and one aquatic therapy expert (n=8). Based on earlier negative feedback about Miro, we used Freeform in this meeting to facilitate collaborative brainstorming and ensure a smoother, more user-friendly experience. This session was focused on summarizing the feedback after the pool session from all the partners (Fig. 4).

**Fig 4.**
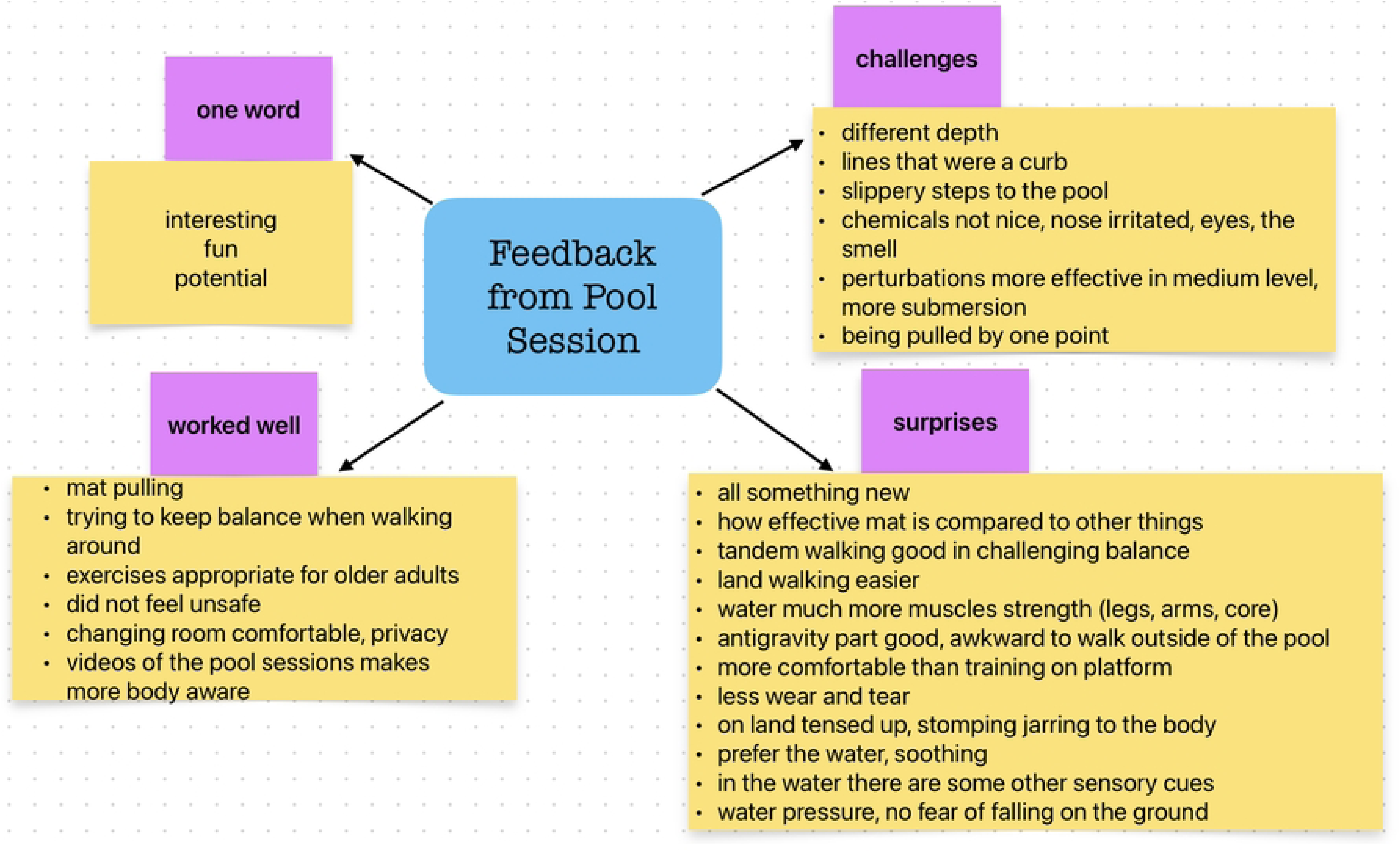
Summarized feedback from the partners after the pool session

Partners provided a range of feedback during the second participatory design session. The water environment was noted to reduce physical strain, increase muscle engagement due to water resistance, and enhance sensory input.

> *“I think they were all more comfortable than the land on the platform that I find the platform is kind of jarring, you know, because you’re anticipating something horrible coming, which doesn’t necessarily happen, but you’re anticipating it” (Partner)*

This statement was made comparing AquaReBal to perturbations provided by a moving platform, experienced by one participant during their participation in an RBT clinical trial. Regarding the AquaReBal exercises, partners felt they were appropriate and safe for older adults. However, they suggested that balance should be tested in future participants both before and after the AquaReBal intervention to accurately assess the effects of the exercises. Feedback from the partners also addressed the equipment used in the exercises.

The mat was considered effective for creating balance challenges, whereas the pool noodles was viewed as less effective. In contrast, the belt was reported as uncomfortable and less effective at generating perturbations.

> *“I didn’t particularly like being pulled from one point and we discussed that a vest might be better because then it’s kind of pulling your whole body, not just one part of your hip” (Partner)*

This statement was from one partner who had felt some discomfort with the hip belt and pointed out that other older adults may experience similar issues. We discussed the possibility of using a vest instead. The pool’s depth presented challenges as the changes of the depth were unclear and confusing. Partners recommended more distinct physical markers or the use of a pool with a gradual slope. Concerns were also raised about the chemicals used in the pool as one partner experienced irritation.

Emotionally, partners described AquaReBal as interesting, enjoyable and promising for future use.

> *“Fun. Interesting. Potential.” (Partner)*

Based on partner input, we collaboratively developed materials for the ‘Introductory package’ to be included in an initial email to new participants (table 1). These included a checklist of items to bring to the sessions, photographs of the pool facilities, a short video demonstrating example exercises. The content was informed by the partners’ own experiences and reflected the resources they felt would have made their initial participation in the study easier.

**Table 1.**
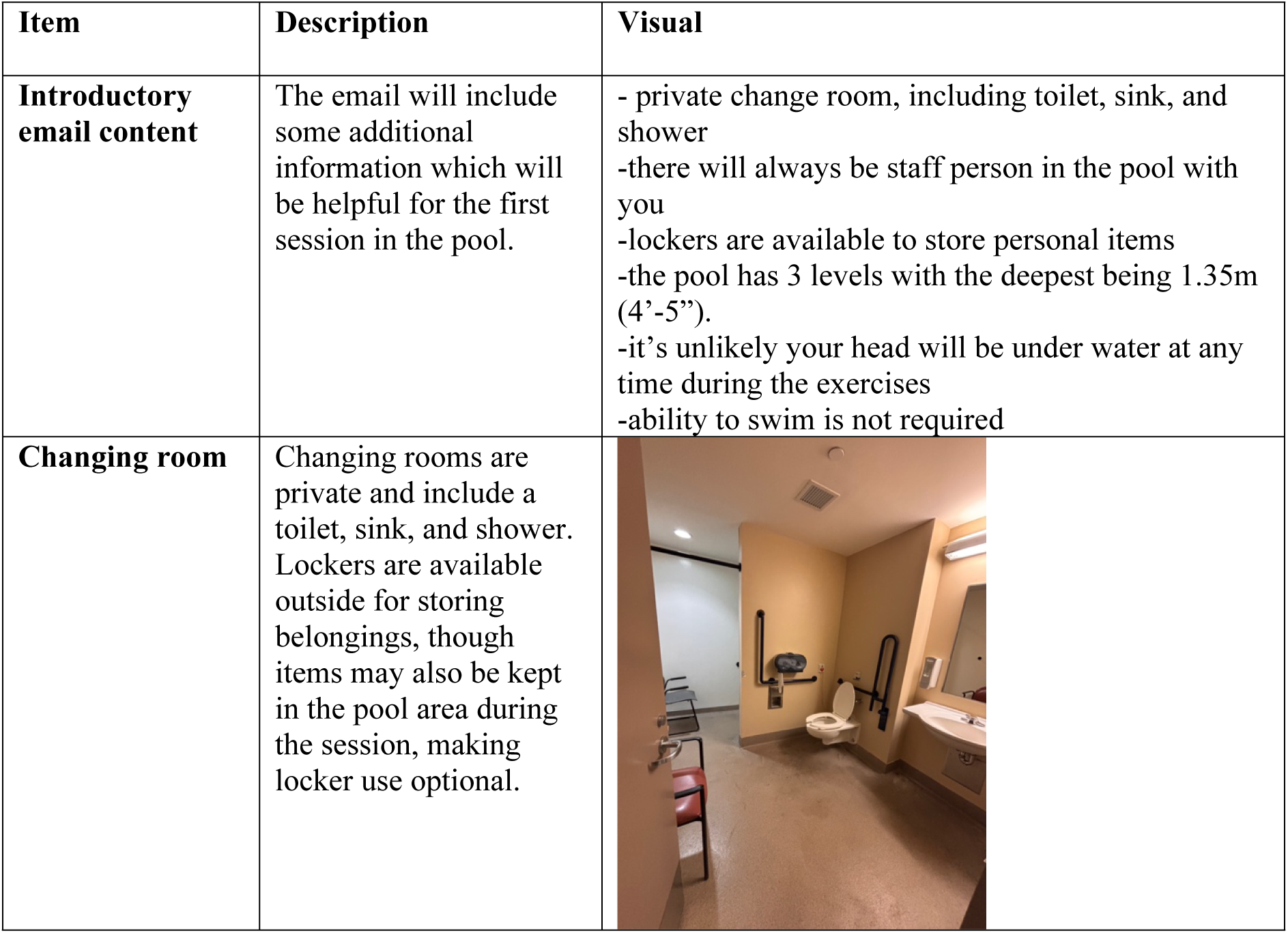
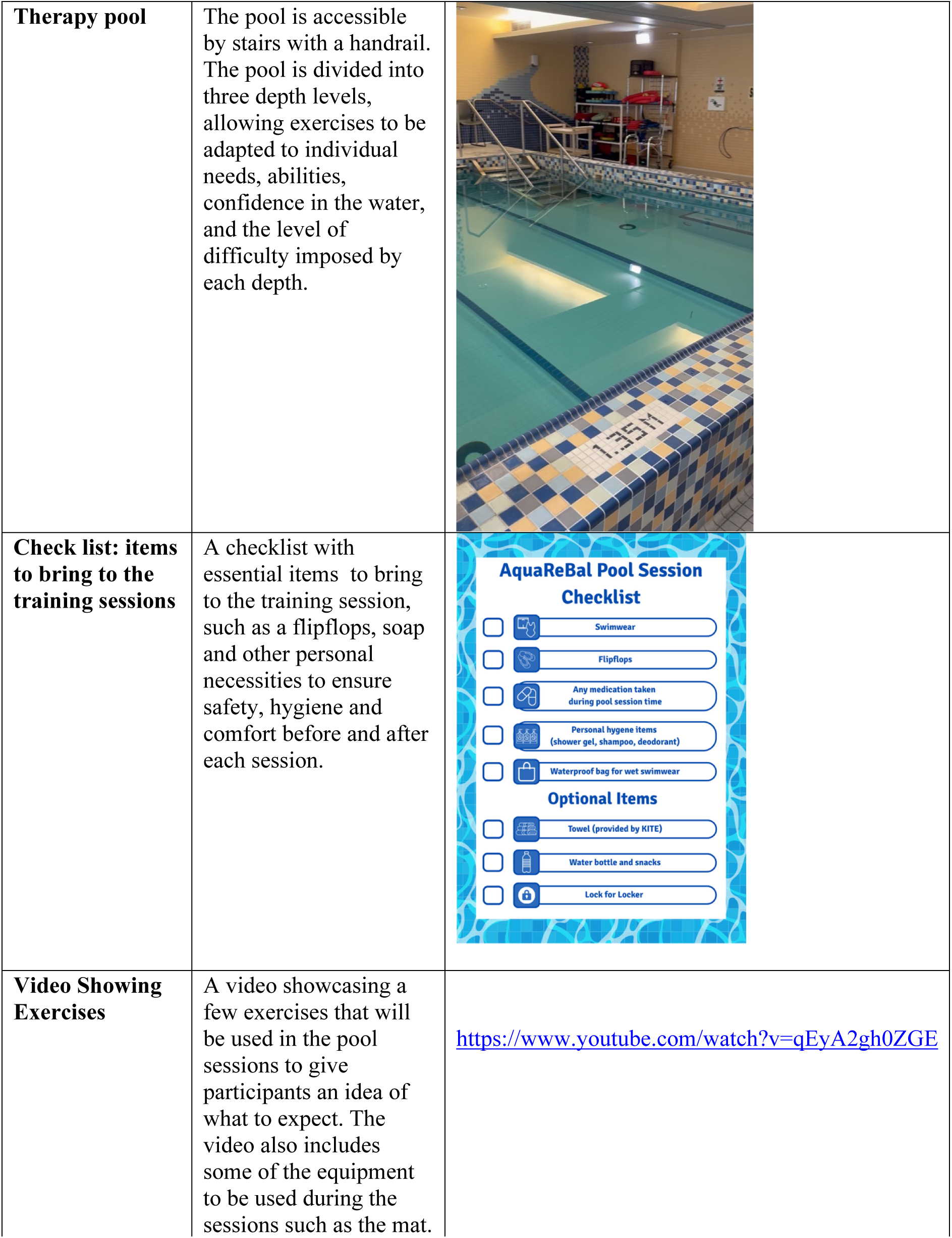
Items included in the introductory package.

| Item | Description | Visual |
| --- | --- | --- |
| **Introductory email content** | The email will include some additional information which will be helpful for the first session in the pool. | - private change room, including toilet, sink, and shower<br>-there will always be staff person in the pool with you<br>-lockers are available to store personal items<br>-the pool has 3 levels with the deepest being 1.35m (4’-5’’).<br>-it’s unlikely your head will be under water at any time during the exercises<br>-ability to swim is not required |
| **Changing room** | Changing rooms are private and include a toilet, sink, and shower. Lockers are available outside for storing belongings, though items may also be kept in the pool area during the session, making locker use optional. |  |
| **Therapy pool** | The pool is accessible by stairs with a handrail. The pool is divided into three depth levels, allowing exercises to be adapted to individual needs, abilities, confidence in the water, and the level of difficulty imposed by each depth. |  |
| **Check list: items to bring to the training sessions** | A checklist with essential items to bring to the training session, such as a flipflops, soap and other personal necessities to ensure safety, hygiene and comfort before and after each session. |  |
| **Video Showing Exercises** | A video showcasing a few exercises that will be used in the pool sessions to give participants an idea of what to expect. The video also includes some of the equipment to be used during the sessions such as the mat. | https://www.youtube.com/watch?v=qEyA2gh0ZGE |

The meeting was evaluated in an anonymous way with a post-meeting survey (n=6) to gather feedback. Two people did not fill in the survey. The meeting was overall assessed positively (4.6/5). People valued the active engagement of meeting attendees, the opportunity to share feedback and reflections, and the focused discussion on aspects of the intervention, such as what worked better or worse for perturbations. The structured format was also appreciated, as it allowed everyone to contribute. Suggestions for improvement included shortening the session, providing discussion questions in advance to allow for more considered responses, and offering a brief overview of study eligibility criteria, goals, and objectives to ensure feedback was well aligned with the research aims. All respondents reported that their ideas were heard and considered. No additional topics or activities were proposed, and overall feedback was highly positive, with one person describing in the comments the experience as “10/10 or 5/5.”

### Internal Team Session 2

During the second internal team session (n=7) the research team reviewed detailed feedback from partners on the aquatic reactive balance exercises, equipment, environmental challenges, and emotional responses. Specific adjustments were discussed, including modifications to perturbation delivery and refinements to session progression to optimize safety and challenge. Outcomes of this session were directly applied to finalize the AquaReBal program ensuring the intervention was responsive to user experiences and ready for implementation.

## Discussion

This study describes the co-design process for developing the AquaReBal program, a novel aquatic RBT intervention for older adults. By integrating evidence from the literature, expert consultation, and direct engagement with older adult partners, the program was designed to be evidence-informed, tailored to the unique characteristics of the aquatic environment and incorporating lived experience.

The co-design approach used in this study not only informed the development of AquaReBal but also highlighted the feasibility of engaging older adults in meaningful ways throughout the research process. Team members and partners reported high satisfaction with participatory sessions, felt their input was valued, and contributed to shaping both the intervention content and delivery. This supports the growing movement toward patient and public involvement in rehabilitation research, which has been associated with improved relevance, uptake, and sustainability of interventions [21].

Building on international guidance for reporting patient and public involvement - the GRIPP2 framework [11], our study illustrates the value of transparent reporting of co-design processes in rehabilitation research. In line with GRIPP2, we documented the aims of involving older adults, the methods of engagement, and the outcomes of involvement.

Contextual factors, such as partners’ previous experience with aquatic exercise and the supportive facilitation of researchers, positively influenced the process. Importantly, the involvement of older adults not only shaped practical elements of AquaReBal but also contributed to theory development by demonstrating how co-design in aquatic environments can extend models of participatory intervention design. Transparent reporting of these processes is critical for building a cumulative evidence base and will support replication and scaling of participatory aquatic interventions in future research [22].

Our findings highlight several key considerations for developing aquatic interventions. First, participants emphasized the importance of safety, comfort, and confidence in the water. This aligns with prior research indicating that fear of falling and environmental barriers are significant determinants of older adults’ participation in aquatic exercise programs [23]. The use of water as a medium for RBT appeared to mitigate some of the physical strain and joint discomfort associated with land-based perturbations, supporting previous evidence that aquatic environments provide a safe and supportive context for balance training in older adults [4]. Considering the main drawbacks of land based RBT is the joint pain and discomfort, this statement supports the use of water environment as more comfortable. Social and relational aspects were consistently identified as motivating factors. Partners highlighted the role of instructors in fostering engagement, enjoyment, and continued participation. This is consistent with research showing that instructor support, feedback, and social interaction can enhance adherence to exercise programs among older adults [24].

Practical and logistical considerations, including equipment, pool layout, and environmental cues, were critical for optimizing safety and usability. Feedback on the hip belt versus vest for perturbations and recommendations for pool depth demonstrate the importance of iterative, participatory design in adapting interventions to the real-world needs of older adults. This reinforces the value of including end-users in program development, ensuring that interventions are acceptable, and aligned with older adults’ preferences [25].

While participants valued the opportunity to shape the intervention, challenges emerged in balancing diverse preferences, reconciling safety concerns with feasibility, and navigating logistical constraints of pool access and equipment availability. Such challenges mirror broader findings in the PPI literature that underline the need to acknowledge both benefits and limitations of involvement rather than presenting idealized accounts [26].

Contextual factors, such as the supportive facilitation provided by researchers, pre-existing trust with partners, and their prior exposure to aquatic exercise, facilitated meaningful engagement. At the same time, process-related factors, including the limited time available for iterative cycles and scheduling constraints, posed barriers to deeper collaboration.

Our work extends existing models of co-design by situating them in an aquatic environment, a context that has been underexplored in rehabilitation research. By embedding PPI from the earliest stages, AquaReBal contributes to the evidence base that co-design can generate not only more acceptable interventions but also novel insights into how balance training is experienced in specific settings.

## Limitations

The number of older adult partners involved in co-design was small and consisted only of women, which may limit the generalizability of the findings. Additionally, partners were relatively active and may not fully represent older adults with higher levels of frailty or functional limitations. Environmental and logistical factors specific to the Toronto Rehabilitation Institute pool may not generalize to other settings. Finally, while participatory design provided rich qualitative insights, the effectiveness of AquaReBal in improving reactive balance control and reducing falls remains to be tested.

## Implications and future directions

The co-design methodology described here can serve as a model for developing other rehabilitation interventions that integrate end-user perspectives, environmental considerations, and evidence-based principles. The findings suggest that AquaReBal has the potential to be a safe, engaging, and adaptable program for older adults. Future research should evaluate the program’s feasibility, acceptability, and preliminary effectiveness in a larger, diverse cohort.

## Conclusions

In conclusion, the creation of AquaReBal protocol demonstrates that co-design in the development of exercise interventions can enhance safety, enjoyment, and relevance, while addressing barriers to participation. The program provides a promising approach for fall prevention, blending evidence-based principles of RBT with the unique advantages of the aquatic environment.

## Acknowledgment

The authors would like to thank all participants for their time and participation in this study. We also acknowledge the support of our research team and collaborating institutions for their valuable contributions throughout the project.

## Ethics approval and consent to participate

The study was approved by the University Health Network Research Ethics Board (study ID: 24-6146).

## Data availability statement

The data that support the findings of this study are available from the corresponding author upon reasonable request.

## References

1. Vaishya R, Vaish A. Falls in Older Adults are Serious. Indian J Orthop. 2020;54(1):69–74. Epub 20200124. doi: 10.1007/s43465-019-00037-x. PubMed PMID: 32257019; PubMed Central PMCID: PMCPMC7093636.

2. Devasahayam AJ, Farwell K, Lim B, Morton A, Fleming N, Jagroop D, et al. The Effect of Reactive Balance Training on Falls in Daily Life: An Updated Systematic Review and Meta-Analysis. Physical therapy. 2023;103(1). doi: 10.1093/ptj/pzac154.

3. Gerards MHG, Sieben J, Marcellis R, de Bie RA, Meijer K, Lenssen AF. Acceptability of a perturbation-based balance training programme for falls prevention in older adults: a qualitative study. BMJ Open. 2022;12(2):e056623. Epub 20220224. doi: 10.1136/bmjopen-2021-056623. PubMed PMID: 35210345; PubMed Central PMCID: PMCPMC8883254.

4. Deng Y, Tang Z, Yang Z, Chai Q, Lu W, Cai Y, et al. Comparing the effects of aquatic-based exercise and land-based exercise on balance in older adults: a systematic review and meta-analysis. Eur Rev Aging Phys Act. 2024;21(1):13. Epub 20240519. doi: 10.1186/s11556-024-00349-4. PubMed PMID: 38764039; PubMed Central PMCID: PMCPMC11102618.

5. Elbar O, Tzedek I, Vered E, Shvarth G, Friger M, Melzer I. A water-based training program that includes perturbation exercises improves speed of voluntary stepping in older adults: a randomized controlled cross-over trial. Arch Gerontol Geriatr. 2013;56(1):134–40. Epub 20120828. doi: 10.1016/j.archger.2012.08.003. PubMed PMID: 22951028.

6. Constantin N, Edward H, Ng H, Radisic A, Yule A, D’Asti A, et al. The use of co-design in developing physical activity interventions for older adults: a scoping review. BMC Geriatr. 2022;22(1):647. Epub 20220808. doi: 10.1186/s12877-022-03345-4. PubMed PMID: 35941570; PubMed Central PMCID: PMCPMC9358386.

7. James H, Buffel T. Co-research with older people: a systematic literature review. Ageing and Society. 2023;43(12):2930–56. Epub 2022/02/10. doi: 10.1017/S0144686X21002014.

8. Corrado AM, Benjamin-Thomas TE, McGrath C, Hand C, Laliberte Rudman D. Participatory Action Research With Older Adults: A Critical Interpretive Synthesis. The Gerontologist. 2020;60(5):e413–e27. doi: 10.1093/geront/gnz080. PubMed PMID: 31264680.

9. Hand C, Keber A, McFarland J, McGrath C, Laliberte Rudman D, Seale L, et al. Neighbourhood-based participatory action research with older adults: Facilitating participation through virtual and remote methods. Methodological Innovations. 2024;17(4):248–60. doi: 10.1177/20597991241292368.

10. McKay H, Nettlefold L, Bauman A, Hoy C, Gray SM, Lau E, et al. Implementation of a co-designed physical activity program for older adults: positive impact when delivered at scale. BMC public health. 2018;18(1):1289. Epub 20181123. doi: 10.1186/s12889-018-6210-2. PubMed PMID: 30470209; PubMed Central PMCID: PMCPMC6251145.

11. Staniszewska S, Brett J, Simera I, Seers K, Mockford C, Goodlad S, et al. GRIPP2 reporting checklists: tools to improve reporting of patient and public involvement in research. BMJ (Clinical research ed). 2017;358:j3453. Epub 20170802. doi: 10.1136/bmj.j3453. PubMed PMID: 28768629; PubMed Central PMCID: PMCPMC5539518.

12. Cusack C, Cohen B, Mignone J, Chartier MJ, Lutfiyya Z. Participatory action as a research method with public health nurses. Journal of advanced nursing. 2018;74(7):1544–53. Epub 20180406. doi: 10.1111/jan.13555. PubMed PMID: 29489024.

13. White GW, Suchowierska M, Campbell M. Developing and systematically implementing participatory action research. Archives of physical medicine and rehabilitation. 2004;85(4 Suppl 2):S3–12. doi: 10.1016/j.apmr.2003.08.109. PubMed PMID: 15083417.

14. Howard Z, Somerville MM. A comparative study of two design charrettes: implications for codesign and participatory action research. CoDesign. 2014;10(1):46–62. doi: 10.1080/15710882.2014.881883.

15. Tremblay MC, Bradette-Laplante M, Bérubé D, Brière É, Moisan N, Niquay D, et al. Engaging indigenous patient partners in patient-oriented research: lessons from a one-year initiative. Res Involv Engagem. 2020;6:44. Epub 20200722. doi: 10.1186/s40900-020-00216-3. PubMed PMID: 32760594; PubMed Central PMCID: PMCPMC7376932.

16. Harrison JD, Auerbach AD, Anderson W, Fagan M, Carnie M, Hanson C, et al. Patient stakeholder engagement in research: A narrative review to describe foundational principles and best practice activities. Health Expect. 2019;22(3):307–16. Epub 20190213. doi: 10.1111/hex.12873. PubMed PMID: 30761699; PubMed Central PMCID: PMCPMC6543160.

17. Leask CF, Sandlund M, Skelton DA, Altenburg TM, Cardon G, Chinapaw MJM, et al. Framework, principles and recommendations for utilising participatory methodologies in the co-creation and evaluation of public health interventions. Res Involv Engagem. 2019;5:2. Epub 20190109. doi: 10.1186/s40900-018-0136-9. PubMed PMID: 30652027; PubMed Central PMCID: PMCPMC6327557.

18. Bazzano AN, Martin J, Hicks E, Faughnan M, Murphy L. Human-centred design in global health: A scoping review of applications and contexts. PloS one. 2017;12(11):e0186744. Epub 20171101. doi: 10.1371/journal.pone.0186744. PubMed PMID: 29091935; PubMed Central PMCID: PMCPMC5665524.

19. Slattery P, Saeri AK, Bragge P. Research co-design in health: a rapid overview of reviews. Health Res Policy Syst. 2020;18(1):17. Epub 20200211. doi: 10.1186/s12961-020-0528-9. PubMed PMID: 32046728; PubMed Central PMCID: PMCPMC7014755.

20. Farlie MK, Keating JL, Molloy E, Bowles KA, Neave B, Yamin J, et al. The Balance Intensity Scales for Therapists and Exercisers Measure Balance Exercise Intensity in Older Adults: Initial Validation Using Rasch Analysis. Physical therapy. 2019;99(10):1394–404. doi: 10.1093/ptj/pzz092. PubMed PMID: 31309981; PubMed Central PMCID: PMCPMC6821236.

21. Brett J, Staniszewska S, Mockford C, Herron-Marx S, Hughes J, Tysall C, et al. A systematic review of the impact of patient and public involvement on service users, researchers and communities. Patient. 2014;7(4):387–95. doi: 10.1007/s40271-014-0065-0. PubMed PMID: 25034612.

22. Brown N. Scope and continuum of participatory research. International Journal of Research & Method in Education. 2022;45(2):200–11. doi: 10.1080/1743727X.2021.1902980.

23. Becker BE. Aquatic therapy: scientific foundations and clinical rehabilitation applications. PM R. 2009;1(9):859–72. doi: 10.1016/j.pmrj.2009.05.017. PubMed PMID: 19769921.

24. Morrison L, McDonough MH, Zimmer C, Din C, Hewson J, Toohey A, et al. Instructor Social Support in the Group Physical Activity Context: Older Participants’ Perspectives. J Aging Phys Act. 2023;31(5):765–75. Epub 20230322. doi: 10.1123/japa.2022-0140. PubMed PMID: 36948211.

25. Concannon TW, Meissner P, Grunbaum JA, McElwee N, Guise JM, Santa J, et al. A new taxonomy for stakeholder engagement in patient-centered outcomes research. J Gen Intern Med. 2012;27(8):985–91. Epub 20120413. doi: 10.1007/s11606-012-2037-1. PubMed PMID: 22528615; PubMed Central PMCID: PMCPMC3403141.

26. Agyei-Manu E, Atkins N, Lee B, Rostron J, Dozier M, Smith M, et al. The benefits, challenges, and best practice for patient and public involvement in evidence synthesis: A systematic review and thematic synthesis. Health Expect. 2023;26(4):1436–52. Epub 20230601. doi: 10.1111/hex.13787. PubMed PMID: 37260191; PubMed Central PMCID: PMCPMC10349234.

